# Social, Financial and Psychological Stress during an Emerging Pandemic: Observations from a Population Web-Based Survey in the acute phase of the COVID-19 pandemic

**DOI:** 10.1101/2020.06.29.20142638

**Authors:** Rebecca Robillard, Mysa Saad, Jodi D. Edwards, Elizaveta Solomonova, Marie-Helene Pennestri, Alexander Daros, Samuel Paul Louis Veissière, Lena Quilty, Karianne Dion, Ashley Nixon, Jennifer L. Phillips, Raj Bhatla, Edward Spilg, Roger Godbout, Bashour Yazji, Cynda Hylton Rushton, Wendy Gifford, Mamta Gautam, Addo Boafo, Tetyana Kendzerska

## Abstract

**Background:** Understanding the multifaceted impacts of the Coronavirus-19 (COVID-19) outbreak as it unfolds is crucial to identify the most critical needs and to inform targeted interventions.

**Methods:** This population survey study presents cohort characteristics and baseline observations linked to the acute-mid phase of the COVID-19 outbreak in terms of perceived threats and concerns, occupational and financial impacts, social impacts and stress as measured by the Cohen’s Perceived Stress Scale (PSS) collected cross-sectionally between April 3 and May 15, 2020. A multivariate linear regression model was used to identify factors associated with stress changes relative to pre-outbreak estimates.

**Findings:** 6,040/6,685 (90·4%) participants filled out at least 1/3 of the survey and were included in the analyses. On average, PSS scores increased from 12·9+6·8 before the outbreak to 14·9+8·3 during the outbreak (p<0·001). The independent factors associated with stress worsening were: having a mental disorder, female sex, having underage children, heavier alcohol consumption, working with the general public, shorter sleep duration, younger age, less time elapsed since the start of the outbreak, lower stress before the outbreak, worse symptoms that could be linked to COVID-19, lower coping skills, worse obsessive-compulsive symptoms related to germs and contamination, personalities loading on extraversion, conscientiousness and neuroticism, left wing political views, worse family relationships, and spending less time exercising and doing artistic activities.

**Interpretation:** Cross-sectional analyses showed a significant increase from average low to moderate stress during the COVID-19 outbreak. Identified modifiable factors associated with an increase in stress may be informative for intervention development.

## INTRODUCTION

An outbreak of Coronavirus Disease (COVID-19), a cluster of acute febrile respiratory illness, was first reported in Wuhan, China, in December 2019.^1^ The World Health Organization declared a pandemic on March 11, 2020, after infections were reported in 110 countries and territories. As of June 4, COVID-19 had spread to 216 countries and territories, infected 6,416,828 individuals, and caused 382,867 deaths worldwide.^2^ This pandemic has created profound economic and social disruption, with the potential for widespread psychological impacts.

Early COVID-19 studies from China, India, Brazil, Paraguay, and the United States indicated high levels of stress with associated sleep problems, poor life satisfaction, and mental illness.^3–7^ As the consequences of this pandemic are likely to be influenced by a range of demographic, occupational, and physical/mental health factors,^6,8,9^ there is a need for comprehensive investigations to identify potential factors modulating psychological responses to this complex situation. Furthermore, most studies to date adopted a broad, representational sampling of adults, but increased efforts to reach individuals at elevated risk for negative outcomes and a lifespan perspective incorporating younger to older age ranges holds particular benefits in informing both prevention and intervention initiatives.

The current manuscript presents the cohort characteristics and baseline observations from an ongoing longitudinal survey launched during the acute phase of the COVID-19 pandemic. Perceived threats and concerns, occupational, financial and social impacts, as well as psychological stress changes relative to retrospective pre-outbreak estimates are reported.

## METHODS

### Study Design

A comprehensive longitudinal online survey was distributed via websites, social media, and multiple organizations and hospitals across Canada. This recruitment strategy (see supplementary section for details) was used to target three core groups: people with chronic mental or physical illnesses, healthcare providers, and the general population. While subsequent reports will focus on specific subgroups, the current report introduces the full cohort.

The sole inclusion criterion was to be 12 years of age and older. The survey was available in English and French, nested in a secured access online platform (www.qualtrics.com) and designed on a decisional tree structure. It included a set of validated questionnaires and custom-built questions pertaining to the pandemic (see supplementary section). Themes covered in the current report include: factors linked to the pandemic (e.g., testing, perceived threat and concerns); occupational and financial life; social life, and psychological stress. Retrospective questions were used to estimate temporal changes from “before the outbreak” (i.e. in the last month before the outbreak) to “during the outbreak” (i.e. in the seven days prior to filling out the survey). The survey was developed and conducted following guidelines from the Checklist for Reporting Results of Internet E-Surveys.^10^ Additional information about the survey and the psychometric properties of validated scales included are outlined in supplemental material.

Electronic informed consent was obtained from each participant. This study was approved by the Clinical Trials Ontario - Qualified Research Ethics Board via the Ottawa Health Science Network (Protocol #2131) and registered at ClinicalTrials.gov (NCT04369690).

### Primary outcome: Psychological stress

Respondents retrospectively assessed their stress levels on the Cohen’s Perceived Stress Scale (PSS)^11^ before and during the outbreak. PSS scores were analyzed continuously (i.e. scale of 0 to 40, estimated minimal clinically important relative change: 28%)^12^ and categorically based on established thresholds: 0 to 13 (low stress), 14 to 26 (moderate stress), and 27 to 40 (high stress).

Factors hypothesized *a priori* to be associated with stress changes were: pre-outbreak stress level, time elapsed since the pandemic declaration by the WHO, age, sex, education level, total family income, employment status, working with the general public, political views, having underage children, having travelled abroad in the past 60 days, index reflective of the number and severity of potential COVID-19 symptoms, the Dimensional Obsessive-Compulsive Scale (DOCS) contamination subscale, Big5 personality subscales, Brief Resilient Coping Scale (BRCS), having a mental disorder, alcohol and drugs use, having a physical condition at risk for COVID-19, sleep duration, quality of family relationships, and amount of time spent outdoors, interacting with other people, following the news on COVID-19, and engaging in physical and artistic activities.

### Analyses

Descriptive statistics were used to characterize survey respondents. To assess changes before and during the outbreak, Chi-squared analyses, paired t-tests/Wilcoxon tests, and McNemar-Bowker tests were used. A repeated measures ANOVA was used to assess the unadjusted cross-sectional temporal evolution of PSS change scores.

Multivariate linear regression was used to identify factors independently associated with PSS changes scores using the “enter” pairwise approach with the predictors listed above. To improve sample homogeneity, this model was run solely on the subgroup of Canadian respondents. A series of multivariate linear models were also run to assess the relation between changes in stress and each independent variable separately while accounting for pre-outbreak PSS scores. Analyses were done using the Statistical Package for Social Sciences (IBM SPSS Statistics for Windows, Version 23·0. Armonk, USA). Details on data cleaning procedures are provided in the supplementary material.

## RESULTS

### Survey and sample characteristics

Between April 3^rd^ and May 15^th^ (i.e. 23 to 65 days after the pandemic declaration by the WHO), 6,685 individuals consented to take part in this study and answered the first survey question. All 6,040 respondents who filled out the minimally sufficient portion of the survey (90·4% of those who answered the first question; see details in supplement) were included in the current report. 81·7% (4,933/6,040) respondents completed the entire survey.

Sample characteristics are presented in Table 1. Respondents ranged between 12 and 83 years old. Most respondents were middle-aged, female, Canadian (mostly from Ontario or Quebec), Caucasian, highly educated, lived in an urban residential area, had children, and were employed with a total yearly family income above $40,000.

**Table 1.** Characteristics of the survey responders at the time of the survey completion

|  | Total n | Missing values<br>% (frequencies) | Mean±SD / % (Frequency) |
| --- | --- | --- | --- |
| Time since outbreak start (days) | 6040 | 0·0 (0) | 50·9±11·7 |
| **<u>General demographics</u>** |  |  |  |
| Age | 6034 | 0·1 (6) | 51·8±17·1 |
| Biological Sex (Females) | 6039 | <0·1 (1) | 70·3% (4248) |
| Gender / Sex Change | 5480 | 9·3 (560) |  |
| Male |  |  | 31·6% (1730) |
| Female |  |  | 67·1% (3676) |
| Transexual |  |  | 0·2% (10) |
| Gender queer or expansive |  |  | 0·9% (50) |
| Other |  |  | 0·3% (14) |
| Current Location | 6005 | 0·6 (35) |  |
| Canada |  |  | 97·3% (5845) |
| US |  |  | 1·3% (79) |
| Others* |  |  | 0·7% (40) |
| France |  |  | 0·4% (26) |
| Australia |  |  | 0·2% (15) |
| Ethnicity | 5577 | 7·7 (463) |  |
| Caucasian |  |  | 86·6% (4832) |
| Others |  |  | 5·6% (311) |
| Asian |  |  | 3·4% (191) |
| First Nation, Metis or Inuk |  |  | 2·1% (115) |
| Arab |  |  | 1·2% (68) |
| Black |  |  | 1·1% (60) |
| Non-Citizen (vs not) | 5634 | 6·7 (406) | 6·1% (343) |
| Political Views (Left-Wing / Right-Wing) | 5167 | 14·5 (873) | 44·8% (2313) / 14·6% (754) |
| Education | 5495 | 0·8 (49) |  |
| University certificate, diploma or degree |  |  | 63·6% (3497) |
| College |  |  | 21·8% (1197) |
| High school |  |  | 14·8% (801) |
| **<u>Socioeconomic, occupational and living situation</u>** |  |  |  |
| Total family income (< \$40K/\$40k to \$100K/>\$100K) | 5601 | 7·3 (439) | 11·1% (624)/ 40·6% (2272)/ 48·3% (2705) |
| Employment status | 5958 | 1·4 (82) |  |
| Unemployed/ Retired / Student |  |  | 12·8% (764)/ 30·6% (1822) / 3·6% (213) |
| Employed |  |  | 53·0% (3159) |
| Having work involves contact with the general public (vs not) | 5779 | 4·3 (261) | 14·3% (826) |
| Dwelling (House / Apartment or Condo) | 5417 | 10·3 (623) | 77·4% (4191) / 22·6% (1226) |
| Living situation (Alone / with another person / with multiple people) | 5606 | 7·2 (434) | 20·0% (1123)/ 44·2% (2478)/ 35·8 (2005) |
| Living area (Rural / Urban) | 5565 | 7·9 (475) | 11·8% (665) / 88·2% (4910) |
| **<u>Health and risks factors</u>** |  |  |  |
| C19 Symptoms index (0-30 scale) | 6040 | 0·0 (0) | 2·1±3·6 |
| Presence of Physical condition at risk for COVID-19† (vs not) | 5629 | 6.8 (411) | 52.1% (2934) |
| Sleep duration (hours; Before the outbreak/ During Outbreak) | 4998 | 17.1 (1030) | 7.3±1.2 / 7.2±1.5 |
| Travelled abroad in last 60 days (vs not) | 5548 | 8.1 (492) | 11.0% (608) |
| **<u>Psychological Domain</u>** |  |  |  |
| PSS scores (0-40 scale; Before the outbreak/ During Outbreak) | 5132 | 15.0 (98) | 12.9±6.8 / 14.9±8.3 |
| DOCS - Contamination (0-20 scale) | 4920 | 18.5 (1120) | 6.1±3.7 |
| Big 5 Subscales (2-10 scale) | 4881 | 19.2 (1161) |  |
| Extraversion |  |  | 6.2±2.1 |
| Agreeableness |  |  | 7.4±1.7 |
| Conscientiousness |  |  | 7.8±1.8 |
| Neuroticism |  |  | 5.6±2.3 |
| Openness to Experiences |  |  | 6.9±1.9 |
| Brief Resilient Coping Scale (4-20 scale) | 4856 | 19.6 (1184) | 14.7±2.9 |
| Mental disorder diagnosis (vs not) | 5607 | 7.2 (433) | 29.0% (1626) |
| **<u>Social Domain</u>** |  |  |  |
| Family Relationship (0-100 scale; Before the outbreak/ During Outbreak) | 5328 | 9.5 (572) | 79.5±19.9 / 74.7±25.4 |
| Has underage children (vs not) | 5731 | 5.1 (309) | 17.2% (985) |
| **<u>Behavioral Domain</u>** |  |  |  |
| Number of alcoholic drinks/week (Before the outbreak/ During Outbreak) | 5557 | 7.9 (476) | 4.1±6.5 / 4.8±6.9 |
| Number of cannabis use/week (Before the outbreak/ During Outbreak) | 5512 | 8.6 (518) | 0.9±5.1 / 1.0±5.1 |
| Spent 30min or less: | 5612 | 7.1 (428) |  |
| Outdoor |  |  | 39.3% (2203) |
| Exercising |  |  | 47.7% (2668) |
| Following C19 news |  |  | 44.0% (2457) |
| Interacting with people in person |  |  | 50.6% (2767) |
| Interacting with people virtually |  |  | 39.5% (2194) |
| Doing an artistic activity |  |  | 75.6% (4155) |

Means, standard deviations (SD), frequencies and percentages (calculated on each item's total sample) for main sample characteristics Location Others: Armenia (n=1), Azerbaijan (n=1), Burkina (n=3), Congo (n=1), Czech Republic (n=1), Denmark (n=1), Germany (n=3), Ireland (n=1), Italy (n=1), Ivory Coast (n=1), Jamaica (n=1), Lebanon (n=1), Malaysia (n=1), Netherlands (n=3), New Zealand (n=1), Pakistan (n=1), Poland (n=1), Romania (n=2), Singapore (n=3), Spain (n=1), Sweden (n=1), United Kingdom (n=8), Vietnam (n=1), Other (n=1); Gender expansive: fluid/non-binary; Alcohol consumption (number of drinks per week); Cannabis consumption (number of times per week), Living area based on postal code. † Physical condition at risk for COVID-19: e.g. respiratory, cardiovascular or autoimmune conditions.

### COVID-19 testing, perceived threats/concerns, and changes relative to before the outbreak

79·3% (4,790/6,040) respondents endorsed at least two symptoms that could be linked to COVID-19. 6·7% (404/6,028) of respondents said they had been tested for COVID-19. Of those, 4·5% (18/404) tested positive and 2·7% (11/404) awaited results. Of those who had not been tested, 4·7% (261/5,580) had contacted public health services to be tested. Within this group, 85·4% (222/260) were declined testing. Rates of declined testing were similar between rural (85·0%, 17/20) and urban areas (86·2%, 193/224; Chi-squared=0·02, p=0·886).

Amongst all respondents, 43·0% (2,505/5,829) estimated that a coronavirus infection would pose high to very high threat to their health and 32·8% (484/5,829) estimated moderate threat. A high to very high threat was estimated by 28·1% (1,589/5,653) for their financial situation, 41·5% (1611/3886) for their jobs or businesses, and 62·8% (3,645/5,802) for their country. Figure 1 shows the degree of concerns related to different secondary effects of the outbreak. Overall, the highest concerns pertained to one’s children or relatives not coping well with the situation, closely followed by being unable to access medications or medical services.

**Figure 1.**
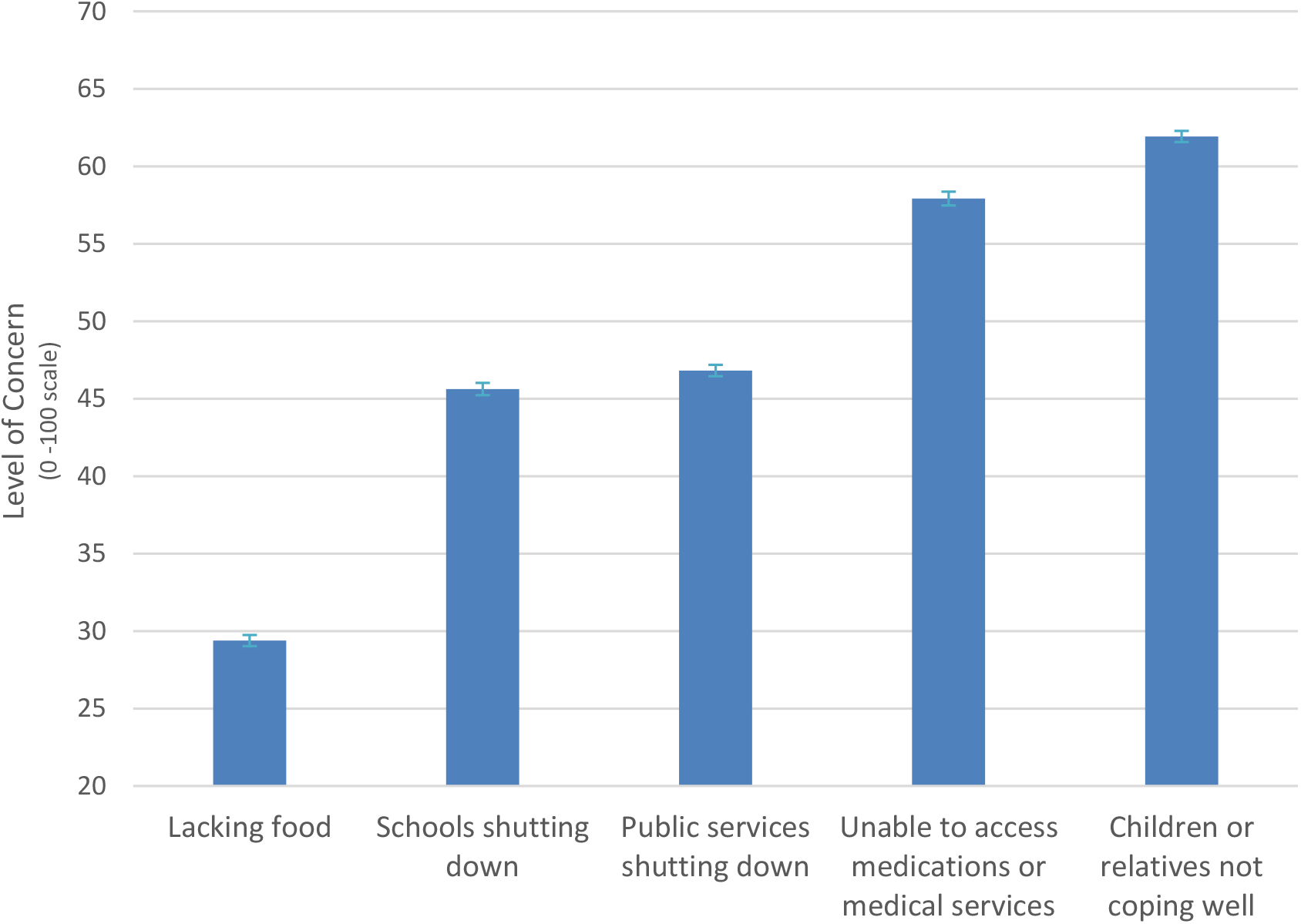
Level of concerns for potential secondary effects of the pandemic Mean level of concerns on a scale ranging from “0-Not concerned at all”, to 50-Neutral” and “100-Very concerned”. Error bars represent standard errors of the mean.

On average, when comparing pre-outbreak estimates and current states: sleep duration shortened (Z=-4·9, p<0·001, r=0·07), family relationships deteriorated (Z=-13·4, p<0·001, r=0·18), and weekly alcohol and cannabis consumption increased (Z=-18·1, p<0·001, r=0·24 and Z=-18·1, p<0·001, r=0·10). Specifically, 10·4% (579/5,563) of the sample over 16 years of age increased their weekly alcohol consumption by five drinks or more.

### Occupational and Financial Impacts

Of the 356 student respondents (Table 1), 84·3% (300/356) reported that their school closed because of the outbreak. Within actively working respondents, 62·8% (2,028/3,228) were working from home, 9·8% (270/3,228) had increased work hours because of the outbreak, and 15·6% (505/3,228) had decreased work hours. 7·9% (254/3,228) underwent a salary decrease due to the outbreak, with an overall median salary reduction of 35% (IQR=50). Of all respondents who were working in the month preceding the outbreak, 11·1% (306/2,764) saw their employment terminated because of the outbreak.

Rates of employment termination due to the outbreak or salary loss exceeding 35% were higher in those with a family income below $40k (41·4%, 82/198) compared to those with higher family income (12·6%, 316/2,503, χ^2^=121·0, p<0·001), in people without a university degree (23·6%, 206/666) compared to in those with a university degree (11·0%; 211/1,913; χ^2^=74·6, p<0·001), and in people with a diagnosis of a mental disorder (16·8%, 137/815) compared to those without (13·5%, 238/1,762; χ^2^=4·9, p=0·027). Rates of employment termination/salary decrease were similar in females versus males (χ^2^=2·3, p=0·132), Caucasians versus other ethnicities (χ^2^=0·9, p=0·335), and people with or without physical illnesses (χ^2^=0·1, p=0·719).

Across the entire sample, 64·5% (3,383/5,243) reported that their expenses had decreased since the start of the outbreak and 15·5% (811/5,243) reported an increase, with a mean estimated rise in health-related expenses of 10·4±20·3%, compared to 29·2±38·0% for food-related expenses.

### Social Life

#### Family and other relationships

Half of parents with underage children (54·0%, 435/806) said that they or their partner were homeschooling. Most respondents estimated that the outbreak was being somewhat disruptive for the management of their work/study and family life (mean rating on a scale from “0 -Very disruptive” to “50-Not different from usual” and “100-Easier than Usual”: 21·6±45·6).

The proportion of respondents interacting with their family more frequently since the start of the outbreak was significantly higher than the proportion of those who were interacting less frequently (p<0·001). The reverse pattern was found for interactions with friends (p<0·001). 40·0% (2,111/5,273) of respondents reported feeling more connected to their family during compared to before the outbreak, while 21·0% (1,107/5,273) felt less connected. This pattern was reversed for connectedness to friends, with 36·2 (1,885/5,210) reporting feeling less connected and 28·3% (1,474/5,210) feeling more connected. On average, relationships ratings with both family and friends during the outbreak significantly deteriorated compared to pre-outbreak estimates (Z=-10·9, p<0·001 and Z=-28·1, p<0·001). Social Distancing

65·8% (3,638/5,530) of respondents were following at least one social distancing guideline at the time of filling out the survey, with 51·6% (2,851/5,530) maintaining a 2 meters distance from others, 46·3% (2,562/5,530) avoiding gatherings in person, 42·5% (2,352/5,530) not using public transport, 37·9% (2,097/5,530) not attending public areas, 35·4% (1,958/5,530) not going out of the home unless they had no choice (e.g. to go to a medical appointment), 29·5% (1,632/5,530) wearing a mask when leaving home, and 17·9% (991/5,530) having food/supplies delivered to their homes. A statistically significant proportion of individuals (between 57·7 to 89·0%) disengaged from some of the social distancing practices that they had initially followed since the start of the outbreak (all p<0·001).

Scores on the UCLA Loneliness Scale were significantly higher in individuals who were avoiding going out of their home (Z=-2·2, p=0·027), living alone (Z=-4·7, p<0·001), younger than 65 years of age (Z=-6·8, p<0·001), diagnosed with a mental disorder (Z=-13·7, p<0·001), or unemployed (Chi-squared=70·0, p<0·001). There was no significant difference in loneliness based on other social distancing practice, sex or whether one worked from home (p>0·050).

### Psychological stress

PSS scores globally increased from 12·9±6·8 before the outbreak to 14·9±8·3 during the outbreak (Z=-22·9, p<0·001, r=0·31), which reflects a transition from low to moderate stress. Rates of individuals with PSS score in the high stress range increased from 3·8% (196/5,132) before the outbreak to 10·2% (535/5,261) during the outbreak (Figure 2). A clinically meaningful increase in stress was noted in 30·3% of respondents, while 10·3% had a clinically meaningful reduction in stress.

**Figure 2.**
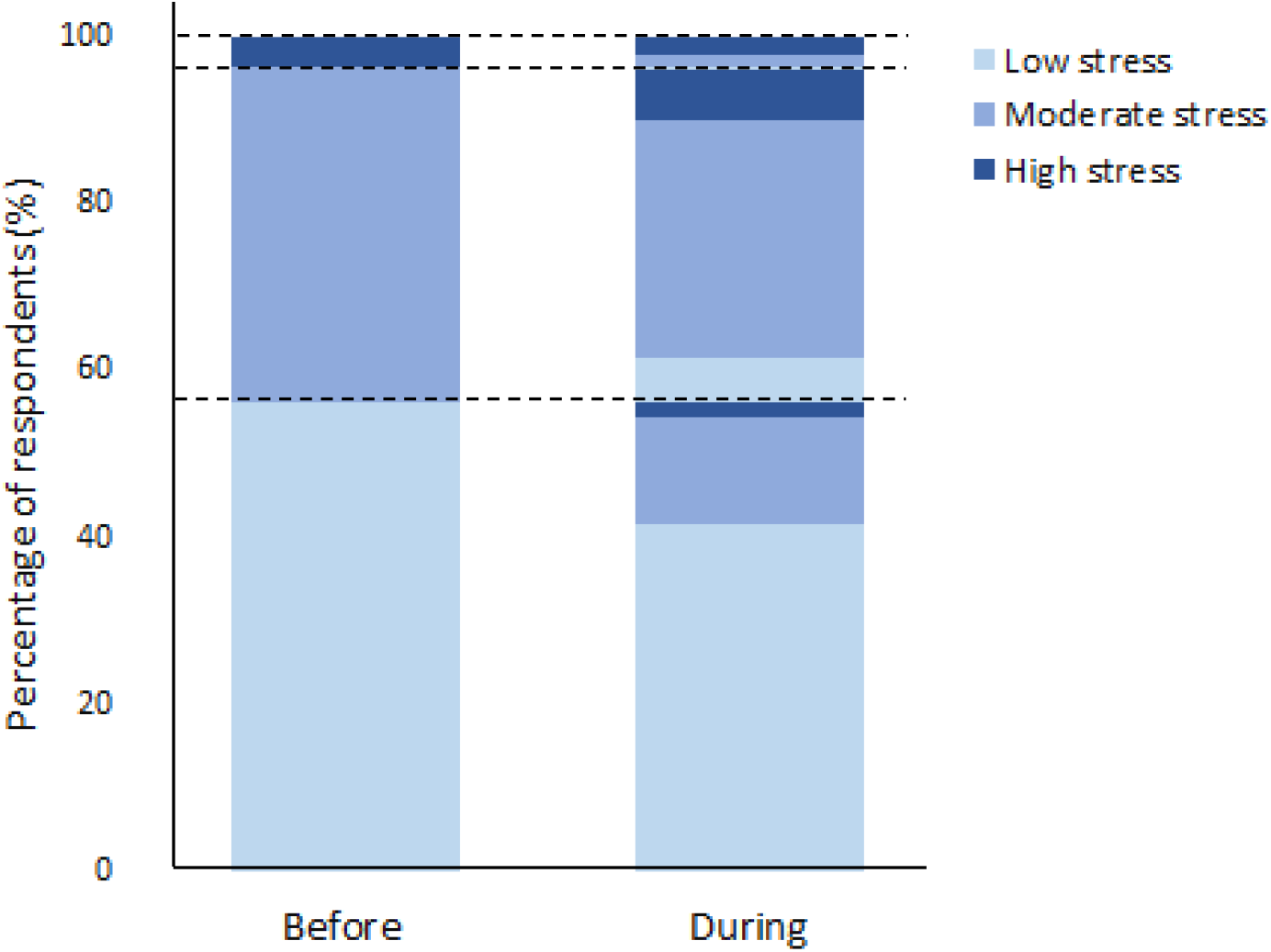
Transitions across stress levels relative to before the outbreak levels Lasagna plot of the percentages (%) of respondents endorsing low, moderate and high stress levels (as per established severity threshold for the Cohen’s Perceived Stress Scale (PSS)) in the retrospective assessment of their stress levels in the month prior to the start of the pandemic (before the outbreak) and in the past 7 days before filling out the survey (i.e. during the outbreak). Dashed lines indicate the transition points between the 3 stress severity ranges. As compared to before the outbreak, 20·8% (1,063/5,103) of respondents had progressed to a higher stress range during the outbreak, and 7·0% (n=355/5,103) of respondents moved to a lower stress range

Over the course of the survey period, there was an overall attenuation of stress worsening on PSS change scores (F (5,5097) =20·07, p<0·001, Figure 3). There was a non-significant reduction in stress worsening between April 3rd and 10th, followed by a plateau which persisted until May 8th, after which there was a significant drop (p≤0·006), compared to all preceding time periods.

**Figure 3.**
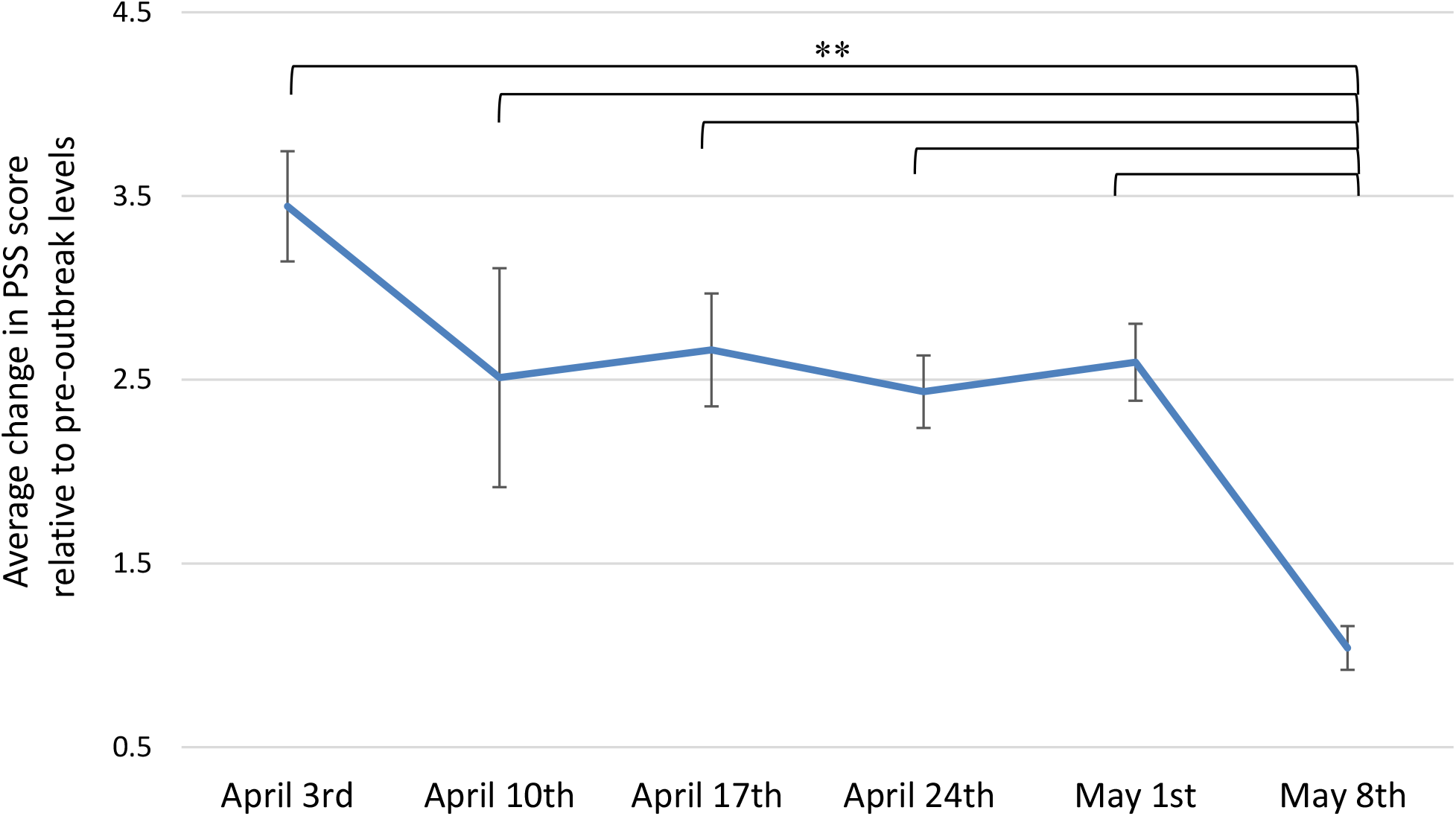
Patterns of stress changes across time Average changes in score on the Cohen’s Perceived Stress Scale (PSS) from pre-outbreak to during the outbreak (i.e. current PSS minus pre-outbreak PSS; higher scores indicating stress worsening) measured cross-sectionally across each time period of survey completion (each comprising 7 days starting on the date of the survey launch). Higher change scores reflect higher stress worsening relative to pre-outbreak stress levels. Error bars indicate the standard error of the mean. Sample sizes for each 7-day time period are as follows: April 3^rd^: n= 516, April 10^th^: n= 135, April 17^th^: n= 453, April 24^th^: n= 1035, May 1^st^: n= 936, May 8^th^: n= 2028. **p<0·001

The following variables were found to be significant independent factors linked to stress worsening in the multivariable linear regression model (Table 2): shorter time elapsed since the start of the outbreak, younger age, female sex, having left wing political views, work involving in-person contact with the general public, having underage children, worse COVID-19 symptoms index, shorter sleep duration, lower PSS scores before the outbreak, higher scores on the DOCS - Contamination subscale and on the extraversion, conscientiousness and neuroticism scales of the Big5, lower BRCS scores, having a mental disorder diagnosis, having had more than five alcoholic drinks in the past week, worse family relationships, and spending less time exercising and doing artistic activities.

**Table 2.** Coefficients of the predictive model for changes in stress

|  | n | Single Predictor Variables |  |  |  |  | Full Model |  |  |  |  |
| --- | --- | --- | --- | --- | --- | --- | --- | --- | --- | --- | --- |
|  |  | B | SE | 95.0% CI |  | p | B | SE | 95.0% CI |  | p |
| Time since outbreak start (7 days) | 5359 | -0.55 | 0.01 | -0.09 | -0.07 | <0.001 | -0.18 | 0.01 | -0.04 | -0.01 | 0.002 |
| **<u>General demographics</u>** |  |  |  |  |  |  |  |  |  |  |  |
| Age (10 years) | 5357 | -0.96 | 0.01 | -0.11 | -0.09 | <0.001 | -0.52 | 0.01 | -0.07 | -0.04 | <0.001 |
| Male sex (female) | 5358 | -2.02 | 0.19 | -2.38 | -1.65 | <0.001 | -0.97 | 0.19 | -1.35 | -0.60 | <0.001 |
| Political Views (vs Center or Others) |  |  |  |  |  |  |  |  |  |  |  |
| Left Wing | 4657 | 0.85 | 0.20 | 0.47 | 1.24 | <0.001 | 0.37 | 0.18 | 0.01 | 0.72 | 0.042 |
| Right Wing | 4657 | 0.21 | 0.28 | -0.34 | 0.75 | 0.457 | 0.31 | 0.24 | -0.17 | 0.79 | 0.206 |
| Education: No university (vs university) | 5327 | -0.20 | 0.18 | -0.55 | 0.16 | 0.277 | -0.22 | 0.19 | -0.59 | 0.14 | 0.230 |
| **<u>Socioeconomic, occupational and living situation</u>** |  |  |  |  |  |  |  |  |  |  |  |
| Total family income (vs >\$100k) | | | | | | | | | | | |
| < \$40k per year | 5009 | 0.72 | 0.31 | 0.12 | 1.33 | 0.019 | 0.30 | 0.18 | -0.05 | 0.65 | 0.094 |
| \$40 to \$100k per year | 5009 | 0.39 | 0.19 | 0.02 | 0.75 | 0.039 | 0.35 | 0.31 | -0.25 | 0.95 | 0.256 |
| Employment status (vs employed): |  |  |  |  |  |  |  |  |  |  |  |
| Unemployed, on leave or student | 5359 | 0.38 | 0.26 | -0.13 | 0.88 | 0.144 | 0.07 | 0.26 | -0.45 | 0.59 | 0.787 |
| Retired | 5359 | -2.37 | 0.19 | -2.75 | -2.00 | <0.001 | -0.15 | 0.25 | -0.64 | 0.34 | 0.544 |
| Work contact with general public (vs not) | 5189 | 1.76 | 0.26 | 1.26 | 2.26 | <0.001 | 0.58 | 0.25 | 0.08 | 1.07 | 0.022 |
| Living in apartment or condo (vs house) | 4858 | 0.36 | 0.21 | -0.05 | 0.77 | 0.089 | -0.10 | 0.21 | -0.50 | 0.31 | 0.631 |
| **<u>Health and risks factors</u>** |  |  |  |  |  |  |  |  |  |  |  |
| C19 Symptoms index (scale from 0 to 30) | 5359 | 0.23 | 0.02 | 0.19 | 0.28 | <0.001 | 0.14 | 0.02 | 0.09 | 0.19 | <0.001 |
| Physical condition at risk† (vs no condition at risk) | 5342 | -0.76 | 0.17 | -1.09 | -0.42 | <0.001 | 0.15 | 0.18 | -0.21 | 0.50 | 0.415 |
| Sleep Duration (hours) | 4804 | -0.59 | 0.06 | -0.1 | -0.48 | <0.001 | -0.53 | 0.05 | -0.64 | -0.42 | <0.001 |
| Travelled abroad in last 60 days (vs no travel) | 4960 | -0.45 | 0.21 | -0.86 | -0.04 | 0.033 | -0.19 | 0.26 | -0.70 | 0.33 | 0.472 |
| **<u>Psychological Domain</u>** |  |  |  |  |  |  |  |  |  |  |  |
| Pre-outbreak PSS (0-40 scale) | 4920 | .. | .. | .. | .. | .. | -0.44 | 0.02 | -0.47 | -0.41 | <0.001 |
| DOCS - Contamination (0-20 scale) | 4717 | 0.47 | 0.02 | 0.43 | 0.52 | <0.001 | 0.38 | 0.02 | 0.34 | 0.42 | <0.001 |
| Big 5 Personality (2-10 scale) |  |  |  |  |  |  |  |  |  |  |  |
| Extraversion | 4680 | 0.15 | 0.04 | 0.07 | 0.23 | <0.001 | 0.13 | 0.04 | 0.06 | 0.21 | 0.001 |
| Agreeableness | 4681 | 0.00 | 0.05 | -0.10 | 0.11 | 0.933 | 0.05 | 0.05 | -0.05 | 0.14 | 0.319 |
| Conscientiousness | 4681 | 0.16 | 0.05 | 0.06 | 0.26 | 0.002 | 0.13 | 0.05 | 0.04 | 0.23 | 0.007 |
| Neuroticism | 4681 | 0.25 | 0.04 | 0.17 | 0.33 | <0.001 | 0.35 | 0.05 | 0.25 | 0.44 | <0.001 |
| Openness to Experiences | 4681 | -0.01 | 0.05 | -0.11 | 0.08 | 0.778 | 0.07 | 0.04 | -0.02 | 0.16 | 0.116 |
| Brief Resilient Coping Scale (4-20 scale) | 1663 | -0.17 | 0.03 | -0.23 | -0.11 | <0.001 | -0.24 | 0.03 | -0.30 | -0.17 | <0.001 |
| Mental disorder diagnosis (vs no diagnosis) | 5326 | 2.34 | 0.20 | 1.95 | 2.74 | <0.001 | 1.14 | 0.20 | 0.74 | 1.54 | <0.001 |
| **<u>Social Domain</u>** |  |  |  |  |  |  |  |  |  |  |  |
| Family Relationship (per 10 units; 0-100 scale) | 5028 | -0.55 | 0.00 | -0.06 | -0.05 | <0.001 | -0.39 | 0.00 | -0.05 | -0.03 | <0.001 |
| Has underage children (vs no underage children) | 5092 | 2.16 | 0.24 | 1.69 | 2.63 | <0.001 | 0.89 | 0.23 | 0.43 | 1.34 | <0.001 |
| **<u>Behavioral Domain</u>** |  |  |  |  |  |  |  |  |  |  |  |
| Weekly alcohol consumption (vs no drinks) |  |  |  |  |  |  |  |  |  |  |  |
| 1 to 5 drinks | 5358 | -0.18 | 0.21 | -0.58 | 0.23 | 0.394 | 0.19 | 0.20 | -0.20 | 0.57 | 0.344 |
| More than 5 drinks | 5358 | 0.15 | 0.21 | -0.27 | 0.56 | 0.490 | 0.61 | 0.20 | 0.21 | 1.01 | 0.003 |
| Weekly cannabis or illicit drugs use (vs no use) | 5312 | 1.13 | 0.26 | 0.63 | 1.63 | <0.001 | 0.45 | 0.25 | -0.03 | 0.93 | 0.066 |
| Spent 30min or less (vs more than 30min): |  |  |  |  |  |  |  |  |  |  |  |
| Outdoor | 5317 | 0.91 | 0.18 | 0.56 | 1.25 | <0.001 | 0.07 | 0.19 | -0.32 | 0.45 | 0.736 |
| Exercising | 5295 | 1.03 | 0.17 | 0.70 | 1.37 | <0.001 | 0.49 | 0.19 | 0.12 | 0.87 | 0.010 |
| Following COVID-19 news | 5296 | -0.25 | 0.17 | -0.59 | 0.08 | 0.141 | -0.24 | 0.17 | -0.57 | 0.09 | 0.155 |
| Social interactions in person | 5201 | 0.14 | 0.17 | -0.20 | 0.48 | 0.406 | 0.21 | 0.16 | -0.11 | 0.53 | 0.205 |
| Social interactions virtually | 5277 | -0.46 | 0.18 | -0.80 | -0.11 | 0.009 | 0.01 | 0.17 | -0.33 | 0.34 | 0.969 |
| Doing an artistic activity | 5210 | 0.16 | 0.20 | -0.23 | 0.56 | 0.421 | 0.50 | 0.19 | 0.12 | 0.88 | 0.010 |

Coefficients parameters for multiple linear regression models including only each single predictors and baseline stress (Left panel) and for the full model (right panel). B: Unstandardized coefficients (calculated per one unit for continuous variables, except for the time elapsed since the start of the outbreak, which was calculated for each 7 days, as well as age and family relationships which were calculated per 10 units). Units (for continuous variables) and reference groups (for categorical variables) are presented in parenthesis in the first column. SE: standard error of B, CI: confidence interval, LL: lower limit, UL: upper limit, Dimensional Obsessive Compulsive Scale (DOCS), Cohen's Perceived Stress Scale (PSS), Liebowitz Social Anxiety Scale (LSAS), Family Relationship rated on scale from "0-Very difficult/conflictual", "50-Neutral" to "100-Excellent". † Physical condition at risk for COVID-19: e.g. respiratory, cardiovascular or autoimmune conditions.

## DISCUSSION

Baseline data from our longitudinal survey in 6,040 respondents suggests that the financial, social and psychological correlates of the COVID-19 outbreak may interact in a complex manner, and that they vary considerably across individuals. While some of our findings echo previous observations, we propose a more comprehensive integrated model of independent factors associated with worse stress responses to this pandemic.

In line with previous polls reporting that many people perceived the COVID-19 pandemic as a greater threat to the economy than to their health,^13^ we observed higher sense of threat related to external/global as opposed to more personal matters. Our observation of concerns about access to medical services are aligned with high rates of potential COVID-19 symptoms with low reported access to testing for COVID-19, a combination which may increase stress.

Consistent with Canadian rates of employment which plummeted by about 11% from February to April 2020,^14^ but lower than the 50% worldwide job losses anticipated by the UN labor agency,^15^ 11% of our respondents lost their job because of the outbreak and an additional 8% underwent salary cuts, with a non-trivial median reduction in salary of 35%. Low income and the lack of a university degree were found to be major risk factors for adverse work and salary outcomes, a phenomenon that may further widen economic disparities. Similarly, reports in the US showed that 40% of people earning $40K or less lost their jobs due to the COVID-19 outbreak and that most of those who kept their job had a university degree.^16^ Importantly, the current study is to our knowledge the first one to identify having a mental disorder as a risk factor for employment termination during the outbreak. The psychological impacts of unemployment are likely to further worsen mental health in these individuals, and they may be at higher risks for subsequent unemployment.^17^ Therefore, this subgroup may face additional challenges not only to cope with the occupational and financial consequences of the pandemic, but also to find work after de-confinement, which highlights potential needs for targeted governmental relief packages and supporting programs to find work.

In line with early COVID-19 reports from China describing major reductions in social contacts beyond the household,^18^ we observed increased interactions with family and decreased interactions with friends, which probably reflect social distancing. This change was accompanied by consistent changes in feelings of connectedness and, paradoxically, by a worsening in relationships quality. Together with previous observations of increased family violence during the pandemic,^19^ this stresses the need to better understand how close proximity in the context of confinement may create family tensions. Only 66% of respondents were following at least one social distancing guideline, a percentage similar to previously reported rates in a previous Canadian poll.^20^ Although the state of emergency still prevailed at the time of the survey, about 60-90% of respondents had been phasing out their social distancing practices. This raises considerable concerns since even a 20% increase in adherence to social distancing can contribute to slow the spread of COVID-19.^21^

We found a significant increase in stress co-occurring with the outbreak, with 30% of individuals undergoing clinically meaningful stress worsening. This is consistent with rates of moderate to severe stress reaching 20 to 27% in Asia, Europe, and Australia.^6,9,22–25^ As anticipated, more acute stress reactions were observed in the earlier phases of the outbreak, with a sharp drop shortly after the mortality peak in Canada was announced. These preliminary observations suggest that although the degree of stress worsening during the outbreak may be phasing out for many individuals, two months after the pandemic declaration, stress levels were not fully back to pre-outbreak levels. This supports the need for the development/promotion of self-help tools for stress management.

Having a current diagnosis of a mental disorder was found to be the strongest independent factor linked to stress worsening, a finding consistent with previous observations about pre-existing psychiatric conditions.^6,9,22–25^ This stresses the importance of further investigation in this group who may require more intensive stress management resources. Poorer coping skills and personality traits loading heavily on extraversion, conscientiousness, and neuroticism were also associated with worse increases in stress. High neuroticism has previously been linked to maladaptive stress coping strategies.^26^ While personalities loading on conscientiousness are usually well-organized, goal-directed and more effective in dealing with stress, the uncertainty associated with this unprecedented outbreak may prevent them from relying on their usual coping strategies, leading to heightened stress. Since extraversion is characterized by a tendency to be active and sociable, social distancing measures probably contributed to worse stress responses in extraverted individuals. Accordingly, a Brazilian Covid-19 survey showed that higher extraversion was associated with lower engagement in social distancing practices, likely reflecting how challenging it is for extraverted individuals to reduce their social proximity.^27^ In line with our finding of an association between left-wing views and stress worsening, a recent Gallup poll in the US^28^ found that liberals (as compared to conservatives) were more likely to worry about worst-case outcomes of the pandemic. The politicization of the crisis and associated media bias (with risk-preventive, pro-lockdown perspectives in the liberal media, and the conservative media appearing to take the crisis less seriously) is one possible explanation for worse pandemic-related distress in liberals.

Our results confirm that several factors previously linked to stress, such as female sex, younger age, having children, and having symptoms that could be linked to COVID-19^6,8,9^ independently contribute to stress worsening. While previous reports highlighted increased risks in healthcare workers,^8^ our findings suggest that this extends to other types of workers physically interacting with the public. Importantly, the current study also identified some modifiable factors that were associated with lower stress responses. For instance, protecting a sufficient period for sleep, minimizing alcohol consumption, promoting better family relationships, exercising, and doing artistic activities may be helpful. Since sleep is thought to contribute to emotional regulation,^29^ attenuating the adverse effects of the pandemic on sleep may enable better coping resources. About 30 minutes of moderate-intensity aerobic exercise three times weekly may also boost mood, reduce psychological distress and decrease symptoms of depression and anxiety.^30^ Planning family activities that may help alleviate tensions and foster more positive relations, as well as creating some time and space for individuals to offset the challenges posed by sustained family proximity may also be relevant to manage stress. Appropriate home-schooling support, as well as better work adaptation for parents may also be required. Increased access to testing is likely to have the collateral effect of attenuating stress levels.

The study has several important limitations. First, generalizability is limited by the dissemination strategy and volunteer bias; although our demographic characteristics are consistent with other published surveys. The length and online nature of the survey may have prevented some individuals from completing it. Recall bias may have affected retrospective estimates of pre-outbreak metrics. Although our multivariate model corrected for this, data collection spanned over a month, a period during which we did observe dynamic changes in stress responses. This study also has several strengths, such as a relatively large sample size, the comprehensive set of factors assessed, and its launch in the early/mid phase of the outbreak.

## CONCLUSION

Baseline data in 6,040 respondents who shared their experiences in the early/mid phase of the COVID-19 pandemic highlighted adverse financial, social and psychological outcomes. Our preliminary findings start to draw a comprehensive model integrating multiple independent factors of the stress responses to this pandemic. Modifiable risk factors identified could inform the development of targeted interventions and support. Populations at risk that should be targeted include: people with pre-existing mental disorders, parents of underage children, people with low income, workers interacting with the general public, people with potential COVID-19 symptoms, and those with sleep disruptions.

## Data Availability

Proposals to access data from this study can be submitted to the corresponding author and may be made available upon data sharing agreement.

## ACKNOWLEDGMENTS

We wish to thank all the participants who gave their time to fill out this extensive survey during this period of turmoil. We also extend our gratitude to the ethics boards who rapidly and diligently provided insights on this project to enable a timely launch, the organizations who helped circulate the survey in their networks, and NIVA inc, for their advice on distribution strategies. We thank the Clinical Investigation Unit at the Ottawa Hospital Research Institute for assistance with participant recruitment.

## AUTHOR’S CONTRIBUTIONS

All co-authors were involved in the following: study conception and design, interpretation of data, revising the manuscript critically for the accuracy and important intellectual content, and final approval of the version to be published. All co-authors are accountable for all aspect of the work in ensuring that questions related to the accuracy or integrity of any part of the work are appropriately investigated and resolved. RR, TK, and JE were additionally involved in the participants’ recruitment as site primary investigators. RR, MS, AN and TK were additionally involved in the following: analyses of data and drafting of the manuscript.

## References

1. Paules CI, Marston HD, Fauci AS. Coronavirus Infections-More Than Just the Common Cold. Vol. 323, Journal of the American Medical Association. American Medical Association; 2020. p. 707–8.

2. World Health Organization (WHO). WHO Coronavirus disease (COVID-19) outbreak situation [Internet].Coronavirus disease (COVID-19) outbreak situation. 2020 [cited 2020 Jun 13]. Available from: https://covid19.who.int/?gclid=CjwKCAjw8pH3BRAXEiwA1pvMsXDoze2QLDa_4WTtExJMku1J3er_GLk-MjRPeOb4_6_ECkdivray6hoCh-oQAvD_BwE

3. Li S, Wang Y, Xue J, Zhao N, Zhu T. The impact of covid-19 epidemic declaration on psychological consequences: A study on active weibo users. Int J Environ Res Public Health. 2020 Mar 2;17(6).

4. Lima CKT, Carvalho PM de M, Lima I de AAS, Nunes JVA de O, Saraiva JS, de Souza RI, et al. The emotional impact of Coronavirus 2019-nCoV (new Coronavirus disease). Vol. 287, Psychiatry Research. Elsevier Ireland Ltd; 2020.

5. Nelson B, Pettitt A, Flannery J, Allen N. Rapid assessment of psychological and epidemiological predictors of COVID-19 concern, financial strain, and health-related behavior change in a large online sample. PsyArXiv Prepr.

6. Qiu J, Shen B, Zhao M, Wang Z, Xie B, Xu Y. A nationwide survey of psychological distress among Chinese people in the COVID-19 epidemic: Implications and policy recommendations. Vol. 33, General Psychiatry. BMJ Publishing Group; 2020.

7. Wang C, Pan R, Wan X, Tan Y, Xu L, Ho CS, et al. Immediate psychological responses and associated factors during the initial stage of the 2019 coronavirus disease (COVID-19) epidemic among the general population in China. Int J Environ Res Public Health. 2020 Mar 1;17(5).

8. Limcaoco RSG, Mateos EM, Fernandez JM, Roncero C. Anxiety, worry and perceived stress in the world due to the COVID-19 pandemic, March 2020. Preliminary results. medRxiv. Apr;

9. Newby J, O’Moore K, Tang S, Christensen H, Faasse K. Acute mental health responses during the COVID-19 pandemic in Australia. medRxiv. May;

10. Eysenbach G. Improving the quality of web surveys: The Checklist for Reporting Results of Internet E-Surveys (CHERRIES). Vol. 6, Journal of Medical Internet Research. Journal of Medical Internet Research; 2004.

11. Cohen S, Kamarck T, Mermelstein R. A global measure of perceived stress. J Health Soc Behav. 1983;24(4):385–96.

12. Eskildsen A, Dalgaard VL, Nielsen KJ, Andersen JH, Zachariae R, Olsen LR, et al. Cross-cultural adaptation and validation of the danish consensus version of the 10-item perceived stress scale. Scand J Work Environ Heal. 2015 Sep 5;41(5):486–90.

13. Lacey N. Public divided on whether isolation, travel bans prevent COVID-19 spread; border closures become more acceptable. Ipsos. 2020 Mar 24;

14. Statistics Canada. Canadian Economic Dashboard and COVID-19 [Internet]. 2020 [cited 2020 Jun 13]. Available from: https://www150.statcan.gc.ca/n1/pub/71-607-x/71-607-x2020009-eng.htm

15. UN labour agency. Nearly half of global workforce at risk as job losses increase due to COVID-19: UN labour agency [Internet]. UN News. 2020 [cited 2020 Jun 13]. Available from: https://news.un.org/en/story/2020/04/1062792

16. Board of Governors of the Federal Reserve System. Report on the Economic Well-Being of U.S. Households in 2018. Washington, DC; 2020 May.

17. Olesen SC, Butterworth P, Leach LS, Kelaher M, Pirkis J. Mental health affects future employment as job loss affects mental health: Findings from a longitudinal population study. BMC Psychiatry. 2013 May 24;13(1):144.

18. Zhang J, Litvinova M, Liang Y, Wang Y, Wang W, Zhao S, et al. Changes in contact patterns shape the dynamics of the COVID-19 outbreak in China. Science (80-). 2020 Apr 29;

19. Humphreys KL, Myint MT, Zeanah CH. Increased Risk for Family Violence During the COVID-19 Pandemic. Pediatrics. 2020 Apr 21;

20. Polls – Research Co. [Internet]. 2020 [cited 2020 Jun 13]. Available from: https://researchco.ca/polls/

21. Ottawa COVID19 Projections [Internet]. 2020 [cited 2020 Jun 13]. Available from: https://613covid.ca/#

22. Casagrande M, Favieri F, Tambelli R, Forte G. The enemy who sealed the world: Effects quarantine due to the COVID-19 on sleep quality, anxiety, and psychological distress in the Italian population. Sleep Med. 2020 May 12;

23. Mazza C, Ricci E, Biondi S, Colasanti M, Ferracuti S, Napoli C, et al. A nationwide survey of psychological distress among italian people during the covid-19 pandemic: Immediate psychological responses and associated factors. Int J Environ Res Public Health. 2020 May 1;17(9).

24. Davico C, Ghiggia A, Marcotulli D, Ricci F, Amianto F, Vitiello B. Psychological Impact of the COVID-19 Pandemic on Adults and Their Children in Italy. SSRN Electron J. 2020 May 6;

25. Moreira PS, Ferreira S, Couto B, Machado-Sousa M, Fernandez M, Raposo-Lima C, et al. Protective elements of mental health status during the COVID-19 outbreak in the Portuguese population. medRxiv. 2020 May 1;

26. Kendler KS, Kuhn J, Prescott CA. The Interrelationship of Neuroticism, Sex, and Stressful Life Events in the Prediction of Episodes of Major Depression. Am J Psychiatry. 2004 Apr 1;161(4):631–6.

27. Carvalho L de F, Pianowski G, Gonçalves AP. Personality differences and COVID-19: are extroversion and conscientiousness personality traits associated with engagement with containment measures? Trends psychiatry Psychother. 2020 Apr 9;

28. McCarthy J. U.S. Coronavirus Concerns Surge, Government Trust Slides. Gallup. 2020;1–9.

29. Gruber R, Cassoff J. The Interplay Between Sleep and Emotion Regulation: Conceptual Framework Empirical Evidence and Future Directions. Curr Psychiatry Rep. 2014;16(11).

30. Paolucci EM, Loukov D, Bowdish DME, Heisz JJ. Exercise reduces depression and inflammation but intensity matters. Biol Psychol. 2018 Mar 1;133:79–84.

